# Chronic Pain, Cannabis Legalization and Cannabis Use Disorder in Veterans Health Administration Patients, 2005 to 2019

**DOI:** 10.1101/2023.07.10.23292453

**Authors:** Deborah S. Hasin, Melanie M. Wall, Dan Alschuler, Zachary L. Mannes, Carol Malte, Mark Olfson, Katherine M. Keyes, Jaimie L. Gradus, Magdalena Cerdá, Charles C. Maynard, Salomeh Keyhani, Silvia S. Martins, David S. Fink, Ofir Livne, Yoanna McDowell, Scott Sherman, Andrew J. Saxon

**Affiliations:** Columbia University Irving Medical Center, 630 West 168th Street, New York, NY 10032, USA; Columbia University Mailman School of Public Health, 722 W 168th St, New York, NY 10032, USA; New York State Psychiatric Institute, 1051 Riverside Dr, New York, NY 10032, USA; Health Services Research & Development (HSR&D) Seattle Center of Innovation for Veteran-Centered and Value-Driven Care, Veterans Affairs Puget Sound Health Care System, 1660 S Columbian Way, Seattle, WA 98108, USA; Center of Excellence in Substance Addiction Treatment and Education, VA Puget Sound Health Care System, 1660 S Columbian Way, Seattle, WA 98108, USA; Boston University School of Public Health, 715 Albany St, Boston, MA 02118, USA; New York University, 50 West 4th Street, New York, NY 10012, USA; University of Washington, 1400 Ne Campus Parkway, Seattle, WA 98195, USA; San Francisco VA Health System, 4150 Clement St, San Francisco, CA 94121, USA; University of California at San Francisco, 505 Parnassus Ave, San Francisco, CA 94143, USA; VA Manhattan Harbor Healthcare, 423 E 23rd St, New York, NY 10010, USA; University of Washington School of Medicine, 1959 NE Pacific St, Seattle, WA 98195, USA

## Abstract

**Background:** The risk for cannabis use disorder (CUD) is elevated among U.S. adults with chronic pain, and CUD rates are disproportionately increasing in this group. Little is known about the role of medical cannabis laws (MCL) and recreational cannabis laws (RCL) in these increases. Among U.S. Veterans Health Administration (VHA) patients, we examined whether MCL and RCL effects on CUD prevalence differed between patients with and without chronic pain.

**Methods:** Patients with ≥1 primary care, emergency, or mental health visit to the VHA and no hospice/palliative care within a given calendar year, 2005-2019 (yearly n=3,234,382 to 4,579,994) were analyzed using VHA electronic health record (EHR) data. To estimate the role of MCL and RCL enactment in the increases in prevalence of diagnosed CUD and whether this differed between patients with and without chronic pain, staggered-adoption difference-in-difference analyses were used, fitting a linear binomial regression model with fixed effects for state, categorical year, time-varying cannabis law status, state-level sociodemographic covariates, a chronic pain indicator, and patient covariates (age group [18-34, 35-64; 65-75], sex, and race and ethnicity). Pain was categorized using an American Pain Society taxonomy of painful medical conditions.

**Outcomes:** In patients with chronic pain, enacting MCL led to a 0·14% (95% CI=0·12%-0·15%) absolute increase in CUD prevalence, with 8·4% of the total increase in CUD prevalence in MCL-enacting states attributable to MCL. Enacting RCL led to a 0·19% (95%CI: 0·16%, 0·22%) absolute increase in CUD prevalence, with 11·5% of the total increase in CUD prevalence in RCL-enacting states attributable to RCL. In patients without chronic pain, enacting MCL and RCL led to smaller absolute increases in CUD prevalence (MCL: 0·037% [95%CI: 0·03, 0·05]; RCL: 0·042% [95%CI: 0·02, 0·06]), with 5·7% and 6·0% of the increases in CUD prevalence attributable to MCL and RCL. Overall, MCL and RCL effects were significantly greater in patients with than without chronic pain. By age, MCL and RCL effects were negligible in patients age 18-34 with and without pain. In patients age 35-64 with and without pain, MCL and RCL effects were significant (p<0.001) but small. In patients age 65-75 with pain, absolute increases were 0·10% in MCL-only states and 0·22% in MCL/RCL states, with 9·3% of the increase in CUD prevalence in MCL-only states attributable to MCL, and 19.4% of the increase in RCL states attributable to RCL. In patients age 35-64 and 65-75, MCL and RCL effects were significantly greater in patients with pain.

**Interpretation:** In patients age 35-75, the role of MCL and RCL in the increasing prevalence of CUD was greater in patients with chronic pain than in those without chronic pain, with particularly pronounced effects in patients with chronic pain age 65-75. Although the VHA offers extensive behavioral and non-opioid pharmaceutical treatments for pain, cannabis may seem a more appealing option given media enthusiasm about cannabis, cannabis commercialization activities, and widespread public beliefs about cannabis efficacy. Cannabis does not have the risk/mortality profile of opioids, but CUD is a clinical condition with considerable impairment and comorbidity. Because cannabis legalization in the U.S. is likely to further increase, increasing CUD prevalence among patients with chronic pain following state legalization is a public health concern. The risk of chronic pain increases as individuals age, and the average age of VHA patients and the U.S. general population is increasing. Therefore, clinical monitoring of cannabis use and discussion of the risk of CUD among patients with chronic pain is warranted, especially among older patients.

**Research in Context:** *Evidence before this study:* Only three studies have examined the role of state medical cannabis laws (MCL) and/or recreational cannabis laws (RCL) in the increasing prevalence of cannabis use disorder (CUD) in U.S. adults, finding significant MCL and RCL effects but with modest effect sizes. Effects of MCL and RCL may vary across important subgroups of the population, including individuals with chronic pain. PubMed was searched by DH for publications on U.S. time trends in cannabis legalization, cannabis use disorders (CUD) and pain from database inception until March 15, 2023, without language restrictions. The following search terms were used: (medical cannabis laws) AND (pain) AND (cannabis use disorder); (recreational cannabis laws) AND (pain) AND (cannabis use disorder); (cannabis laws) AND (pain) AND (cannabis use disorder). Only one study was found that had CUD as an outcome, and this study used cross-sectional data from a single year, which cannot be used to determine trends over time. Therefore, evidence has been lacking on whether the role of state medical and recreational cannabis legalization in the increasing US adult prevalence of CUD differed by chronic pain status.

*Added value of this study:* To our knowledge, this is the first study to examine whether the effects of state MCL and RCL on the nationally increasing U.S. rates of adult cannabis use disorder differ by whether individuals experience chronic pain or not. Using electronic medical record data from patients in the Veterans Health Administration (VHA) that included extensive information on medical conditions associated with chronic pain, the study showed that the effects of MCL and RCL on the prevalence of CUD were stronger among individuals with chronic pain age 35-64 and 65-75, an effect that was particularly pronounced in older patients ages 65-75.

*Implications of all the available evidence:* MCL and RCL are likely to influence the prevalence of CUD through commercialization that increases availability and portrays cannabis use as ‘normal’ and safe, thereby decreasing perception of cannabis risk. In patients with pain, the overall U.S. decline in prescribed opioids may also have contributed to MCL and RCL effects, leading to substitution of cannabis use that expanded the pool of individuals vulnerable to CUD. The VHA offers extensive non-opioid pain programs. However, positive media reports on cannabis, positive online “information” that can sometimes be misleading, and increasing popular beliefs that cannabis is a useful prevention and treatment agent may make cannabis seem preferable to the evidence-based treatments that the VHA offers, and also as an easily accessible option among those not connected to a healthcare system, who may face more barriers than VHA patients in accessing non-opioid pain management. When developing cannabis legislation, unintended consequences should be considered, including increased risk of CUD in large vulnerable subgroups of the population.

## INTRODUCTION

The legal status of cannabis varies across countries and time periods. In the United Kingdom, cannabis use is illegal, with stricter legal penalties under discussion^1^. In contrast, Canada legalized cannabis nationally for medical and recreational use in 2001 and 2018, respectively, with legislation that emphased public health concerns^2^. In the United States, legalization has occurred on a state-by state basis: since 1996, 39 U.S. states enacted medical cannabis laws (MCL), and since 2012, 21 states enacted recreational cannabis laws (RCL). Much of the U.S. legislation has been industry-friendly rather than public health focused^3^.

While many individuals can use cannabis without harm, 20%-33% of those who use cannabis develop cannabis use disorder (CUD),^4^ a symptomatic condition with impaired functioning.^5-7^ Over the last two decades, the proportion of Americans favoring cannabis legalization has increased to 68%, the perception that cannabis use is risky has decreased^8, 9^, and rates of U.S. adult cannabis use^8, 10-12^ and CUD have increased.^10, 11, 13-15^ Adjusted for nationally increasing trends, MCL and RCL have been shown to play a role in the increasing prevalence of cannabis use.^16-21^ However, fewer similarly-adjusted national studies examined MCL and RCL effects on trends in the prevalence of CUD, two using the same survey data from U.S. general population^22-24^ and one using data from Veterans Health Administration (VHA) patients.^25^ These studies indicate that MCL and RCL played a role in the increasing prevalence of CUD,^22, 23, 25^ but that their effects were modest. The modest-sized effects could occur if MCL and RCL effects differ across subgroups, obscuring important differences when groups are combined. Specifically, studies have not investigated whether state MCL or RCL effects differ between individuals with and without chronic pain.

Chronic pain, (i.e., pain lasting ≥3 months^26^) is an increasing U.S. health problem.^27-30^ Veterans have high rates of chronic pain.^28, 31^ Pain and CUD are associated.^32,33^ The prevalence of CUD diagnoses has increased disproportionately in VHA patients with chronic pain, particularly older patients,^34^ possibly due to increasing use of cannabis to manage pain,^35^ expanding the pool of individuals using cannabis who are vulnerable to CUD. Most U.S. adults, particularly in MCL and RCL states, believe that cannabis is effective for treating pain^36^ despite inconclusive evidence^37-39^, and pain is the most common qualifying condition for medical cannabis.^40^ Given high rates of pain in VHA patients, the association of pain with CUD, and more prevalent beliefs that cannabis is useful for pain in MCL and RCL states, we hypothesized that MCL and/or RCL would play a greater role in the national increases in rates of CUD in VHA patients with chronic pain than in other patients.

The VHA is the largest integrated U.S. healthcare system^41, 42^ We leveraged VHA electronic health record (EHR) data from 2005-2019 to investigate whether the role of MCL and RCL enactment in the increases in rates of CUD diagnoses differed between patients with and without chronic pain. Because MCL and RCL have stronger effects in older adults^16, 22, 25^ and disproportionate increases in CUD are greatest in older patients with pain,^34^ we also examined results by age group.

## METHODS

Data were obtained through the VHA Corporate Data Warehouse, a data repository for care provided at VHA facilities or paid by the VHA. Veterans 18-75 years with ≥1 VHA primary care, emergency department, or mental health visit in a given calendar year were included, except those in hospice/palliative care or outside the 50 states or Washington DC. Resulting n’s of 3,234,382 to 4,579,994 patients each year were used to create 15 datasets, one for each year, 2005-2019 (see E-Table 1 for yearly ns). New York State Psychiatric Institute, VA Puget Sound and VA New York Harbor Healthcare Systems Institutional Review Boards granted waivers/exemptions of informed consent.

**Table 1.** Adjusted^a^ CUD prevalence in VHA patients in 2005 and 2019, by pain and enacted state law status as of 2019, and absolute change over time, overall and by age group

|  | CUD prevalence<sup>a</sup><br>% |  | Absolute<br>Change | CUD prevalence<sup>a</sup><br>% |  | Absolute<br>Change |
| --- | --- | --- | --- | --- | --- | --- |
| State legal status by 2019 | 2005 | 2019 | % | 2005 | 2019 | % |
|  | **Patients with Chronic Pain** |  |  | **Patients without Chronic Pain** |  |  |
| **Full Sample, Age ≥18** | N = 1,282,021 | N = 2,493,187 |  | N = 1,952,361 | N = 2,086,807 |  |
| No-CL (17 states) | 1.76 | 2.92 | 1.17 | 1.01 | 1.53 | 0.52 |
| MCL-Only (22 states) | 1.76 | 3.37 | 1.61<sup>b</sup> | 1.01 | 1.66 | 0.65<sup>b</sup> |
| MCL/RCL (11 states and DC) | 1.77 | 3.41 | 1.64<sup>c</sup> | 1.03 | 1.73 | 0.70<sup>c</sup> |
| **Age 18-34** | N = 60,101 | N = 221,418 |  | N = 122,708 | N = 261,075 |  |
| No-CL (17 states by 2019) | 1.96 | 5.61 | 3.64 | 1.24 | 3.53 | 2.29 |
| MCL-Only (23 states by 2019)<sup>d</sup> | 2.21 | 6.10 | 3.89<sup>b</sup> | 1.56 | 3.65 | 2.09<sup>b</sup> |
| MCL/RCL (10 states and DC by 2019)<sup>d</sup> | 2.53 | 6.65 | 4.12<sup>c</sup> | 1.59 | 4.02 | 2.43<sup>c</sup> |
| **Age 35-64** | N = 868,355 | N = 1,295,657 |  | N = 1,125,137 | N = 939,778 |  |
| No-CL (17 states by 2019) | 1.57 | 3.49 | 1.92 | 1.00 | 1.89 | 0.89 |
| MCL-Only (23 states by 2019)<sup>d</sup> | 1.69 | 4.26 | 2.57<sup>b</sup> | 1.22 | 2.16 | 0.94<sup>b</sup> |
| MCL/RCL (10 states and DC by 2019)<sup>d</sup> | 1.91 | 4.36 | 2.45<sup>c</sup> | 1.37 | 2.24 | 0.87<sup>c</sup> |
| **Age 65-75** | N = 353,565 | N = 976,112 |  | N = 704,516 | N = 885,954 |  |
| No-CL (17 states by 2019) | 0.48 | 1.34 | 0.86 | 0.18 | 0.56 | 0.38 |
| MCL-Only (23 states by 2019)<sup>d</sup> | 0.52 | 1.62 | 1.10<sup>b</sup> | 0.21 | 0.64 | 0.42<sup>b</sup> |
| MCL/RCL (10 states and DC by 2019)<sup>d</sup> | 0.48 | 1.63 | 1.16<sup>c</sup> | 0.21 | 0.68 | 0.47<sup>c</sup> |

<sup>a</sup> Adjusted for categorical age, sex, race/ethnicity, all age\*race/ethnicity\*sex interactions, state-level median income and percentages: male, Hispanic, non-Hispanic White, non-Hispanic Black, poverty, age 18+, unemployed.
<sup>b</sup> Value used as denominator to determine % of overall increase attributable to MCL enactment based on DiD estimates of MCL effect (see Table 2)
<sup>c</sup> Value used as denominator to determine % of overall increase attributable to RCL enactment based on DiD estimates of RCL effect (see Table 2)
CL=cannabis law; MCL=medical cannabis law; RCL=recreational cannabis law; DC=Washington DC

### Choice of Primary Measure

The primary outcome was a clinician-assigned CUD diagnosis at ≥1 outpatient or inpatient encounter within a calendar year; ICD-9-CM (2005-2015: 305·2X, abuse; 304·3X, dependence), and ICD-10-CM (2016-2019: F12·1X, abuse; F12·2X, dependence), excluding CUD in remission and unspecified cannabis use codes.

As previously described,^34^ patients were classified with chronic pain using an American Pain Society taxonomy and ICD-9-CM-ICD-10-CM crosswalk to identify eleven clusters of chronic pain conditions^43^: back; neck; limb/extremity/joint pain and non-systemic non-inflammatory arthritic disorders; fibromyalgia; headache/migraine; orofacial/ear/temporomandibular disorders; abdominal/bowel disorders; urogenital/pelvic/menstrual disorders; neuropathy; systemic disorders; and other painful conditions. The ICD codes are available elsewhere.^44^ We excluded cancer-related and acute pain, e.g., fractures.^34^ We required diagnoses for specific conditions from ≥2 outpatient visits or ≥1 inpatient hospitalization within each study year. A binary variable was created indicating any chronic pain condition each year from 2005-2019.

Primary exposures were state cannabis laws: state-year variables indicating state MCL and/or RCL enactment. Patient state-of-residence was indicated by last healthcare encounter for each year. States were categorized each year as No-CL, MCL-only, and MCL/RCL. For sensitivity analysis involving state legal dispensaries, which can occur subsequent to MCL or RCL enactment, we used RAND-USC OPTIC marijuana policy data^45^ to create state-year variables indicating the years that legally-protected dispensaries were operational.

Age-group definitions followed previous studies: 18-34, 35-64, and 65-75 years.^15, 25, 34^ Patients ≥76 years were excluded because of their very low CUD prevalence.^25^ Control covariates included age, sex, race and ethnicity (non-Hispanic White, non-Hispanic Black, Hispanic, other/multiple, unknown). Time-varying yearly state control covariates included state/year rates from American Community Survey data: % male; non-Hispanic white; non-Hispanic Black; Hispanic; ≥18 years of age; unemployed; below poverty threshold; and yearly median household income. One-year^46^ and 5-year^47^ estimates were used for 2005-2008 and 2009-2019, respectively, using R tidycensus.^48^

### Statistical Analysis

Initial analyses of diagnosed CUD prevalence across years 2005 to 2019 were grouped by state law status in 2019: (1) No-CL; (2) MCL-only; and (3) MCL/RCL, stratified by chronic pain. Adjusted prevalence estimates were obtained from a linear binomial regression model interacting state law status in 2019 by year by chronic pain, controlling for age, sex, race, ethnicity, and time-varying state covariates. Additionally, models were run stratified by age group.

The effects of MCL and RCL enactment were estimated with the staggered-adoption difference-in-difference (DiD) model.^49^ This model uses each state that enacts a law as its own control by comparing aggregated post-law years to pre-law years while controlling for historical trends using data from all other states that did not enact the respective law in contemporaneous years. A time-varying indicator was constructed for each state-year indicating No-CL, MCL-only, or MCL/RCL for that year. DiD estimates and 95% confidence interval (CI) for MCL-only and MCL/RCL effects were obtained from a linear binomial regression model with fixed effects for state, categorical year, time-varying law status, and a chronic pain indicator, controlling for age, sex, race, ethnicity, and time-varying state-level covariates. Resulting DiD estimates include the effect of a state moving from No-CL to MCL-only and from MCL-only to MCL/RCL among patients with and without pain. Differences in DiD effects between patients with and without pain were determined by the confidence intervals around the DiD effects for both groups; non-overlapping CIs indicated that the DiD effects for the two groups differed significantly. Additional DiD models were run stratified by age group (18-34, 35-64, 65-75), adjusting for within-group continuous age.

As noted previously, the ICD-9-CM to ICD-10-CM transition resulted in a slight downward shift in CUD prevalence in 2015 across the VHA and all states.^25, 50^ DiD estimates take this into account by utilizing states that had not yet passed MCL or RCL (which experienced the ICD transition at the same time) as contemporaneous secular controls.

Following previous procedures,^25^ we illustrate the magnitude of the DiD estimates relative to the overall increases in CUD prevalence (i.e., the amount of change attributable to the laws) by dividing the DiD estimates by the absolute changes between 2005 and 2019 in the states with the respective laws by 2019. To explore whether law effects differed by earlier or later enactment, we produced state-specific DiD estimates using interaction terms between state and time-varying no-CL/MCL-only/RCL status.

Sensitivity analyses used similar methods. We examined legalized dispensaries by replacing state/year no-CL/MCL-only/RCL variables with the year dispensaries were first operational.^45^ We examined lagged MCL-only/RCL effects by replacing MCL-only/RCL state/year variables with 1-year post-enactment dates; this lag maximized the number of included RCL states.

## RESULTS

### Demographic characteristics

E-Table 2 shows patient demographics. The mean age of patients with and without chronic pain ranged from 57·19 (SD=13·78) to 58·33 (SD=12·59) years. Over 90% were male, and most were White.

**Table 2.** MCL and RCL^a^ enactment and changes in CUD prevalence in VHA patients with and without pain: Difference-in-difference (DiD)^b^ estimates using data across all years, 2005 – 2019, overall and by age group

CL=cannabis law; MCL=medical cannabis law; RCL=recreational cannabis law; DC=Washington DC
|  |  |  |  |  |  |
| --- | --- | --- | --- | --- | --- |
|  | Model-based DiD law effect<sup>c</sup><br>%<br>(p-value)<br>95% CI | % of absolute change<br>attributable to law<sup>c</sup> | Model-based DiD law effect<sup>c</sup><br>%<br>(p-value)<br>95% CI | % of absolute change<br>attributable to law<sup>c</sup> | Comparison of DiD effect between<br>Patients<br>With and Without Chronic Pain |
| **Full Sample, Age ≥18** | **Patients With Chronic Pain** |  | **Patients Without Chronic Pain** |  |  |
| Effect of change from no-CL to MCL-only<sup>d</sup> | 0.135<br>(<.001)<br>0.118, 0.153 | 8.4% | 0.037<br>(<.001)<br>0.027, 0.048 | 5.7% | P<0.01 |
| Effect of change from MCL-only to RCL/MCL<sup>d</sup> | 0.188<br>(<.001)<br>0.160, 0.217 | 11.5% | 0.042<br>(<.001)<br>0.023, 0.060 | 6.0% | P<0.01 |
| **Age 18-34** |  |  |  |  |  |
| Effect of change from no-CL to MCL-only<sup>d</sup> | 0.133<br>(0.012)<br>0.029, 0.238 | 3.4% | 0.028<br>(0.377)<br>-0.035, 0.091 | 1.4% | p>0.01 |
| Effect of change from MCL-only to RCL/MCL<sup>d</sup> | -0.053<br>(0.453)<br>-0.192, 0.085 | -1.3% | 0.033<br>(0.441)<br>-0.052, 0.118 | 1.4% | p>0.01 |
| **Age 35-64** |  |  |  |  |  |
| Effect of change from no-CL to MCL-only<sup>d</sup> | 0.203<br>(<.001)<br>0.170, 0.236 | 7.9% | 0.076<br>(<.001)<br>0.053, 0.100 | 8.1% | P<0.01 |
| Effect of change from MCL-only to RCL/MCL<sup>d</sup> | 0.142<br>(<.001)<br>0.093, 0.192 | 5.8% | -0.072<br>(<.001)<br>-0.107, -0.037 | -8.2% | P<0.01 |
| **Age 65-75** |  |  |  |  |  |
| Effect of change from no-CL to MCL-only<sup>d</sup> | 0.102<br>(<.001)<br>0.084, 0.121 | 9.3% | 0.029<br>(<.001)<br>0.017, 0.041 | 6.9% | P<0.01 |
| Effect of change from MCL-only to RCL/MCL<sup>d</sup> | 0.224<br>(<.001)<br>0.191, 0.258 | 19.4% | 0.075<br>(<.001)<br>0.055, 0.096 | 16.1% | P<0.01 |
| <sup>a</sup> MCL=medical cannabis law; RCL=recreational cannabis law.<br><sup>b</sup> Staggered-Adoption Difference-in-Difference (DID) regression model<sup>49</sup>, adjusted for categorical age, sex, race/ethnicity, all age*race/ethnicity*sex interactions, state-level median income and percentages: male, Hispanic, non-Hispanic White, non-Hispanic Black, poverty, age 18+, unemployed. Model estimated effects represent absolute increase (positive values) or decrease (negative values) in CUD prevalence due to law enactment. Confidence intervals not including 0.0 indicate significant changes. The DiD model compares the years after enactment (up to 2019 or until the next law change) in each state to the years before enactment (since 2005 or the previous law change) in the same state, and controls for contemporaneous trends in other states that have not yet passed the respective law.<br><sup>c</sup> 22 states and DC made a change from no-CL to MCL-only during the period from 2005-2019; 11 states and DC made a change from MCL-only to RCL/MCL during the period. Note, 3 of these states and DC made both changes between 2005-2019 (i.e., from no-CL to MCL-only and then later to RCL/MCL), hence contributing information to both effects. There were 20 states (3 with MCL-only and 17 with no-CL in 2019) that made no law changes between 2005-2019; in the DID model, they contribute to background secular trends.<br><sup>d</sup> DiD estimate divided by absolute change across period as shown in Table 1.<br><sup>e</sup> Based on non-overlapping confidence intervals of the two DID effects, one in the group with chronic pain and the other in the group without chronic pain |  |  |  |  |  |

\* Adjusted for categorical age, sex, race/ethnicity, all age\*race/ethnicity\*sex interactions, state-level median income and percentages: male, Hispanic, non-Hispanic White, non-Hispanic Black, poverty, age 18+, unemployed.
\* In states that changed to MCL and RCL during the period, the MCL/RCL effect plotted is compared to No-CL for comparison with the MCL vs No-CL effect.

### CUD prevalence trends, 2005-2019, in states grouped by 2019 cannabis law status

Covariate-adjusted CUD prevalence trends in the no-CL, MCL-only, and MCL/RCL states (defined by their 2019 status) are shown in Table 1 and Figures 1·A and 1·B (patients with and without chronic pain, respectively). Among patients with chronic pain, CUD prevalence in no-CL, MCL-only, and MCL/RCL states increased from 1·76% to 2·92% in no-CL states, from 1·76% to 3·37% in MCL states, and from 1·77% to 3·41% in MCL/RCL states, thus increasing 1·17%, 1·61% and 1·64% in no-CL, MCL-only, and MCL/RCL states. Among patients without chronic pain, CUD prevalence increased from 1·01% to 1·53% in no-CL states, from 1·01% to 1·66% in MCL-only states, and from 1·03% to 1·73% in RCL states, thus increasing 0·52%, 0·65% and 0·70% in no-CL, MCL-only, and MCL/RCL states, respectively.

**Figure 1.**
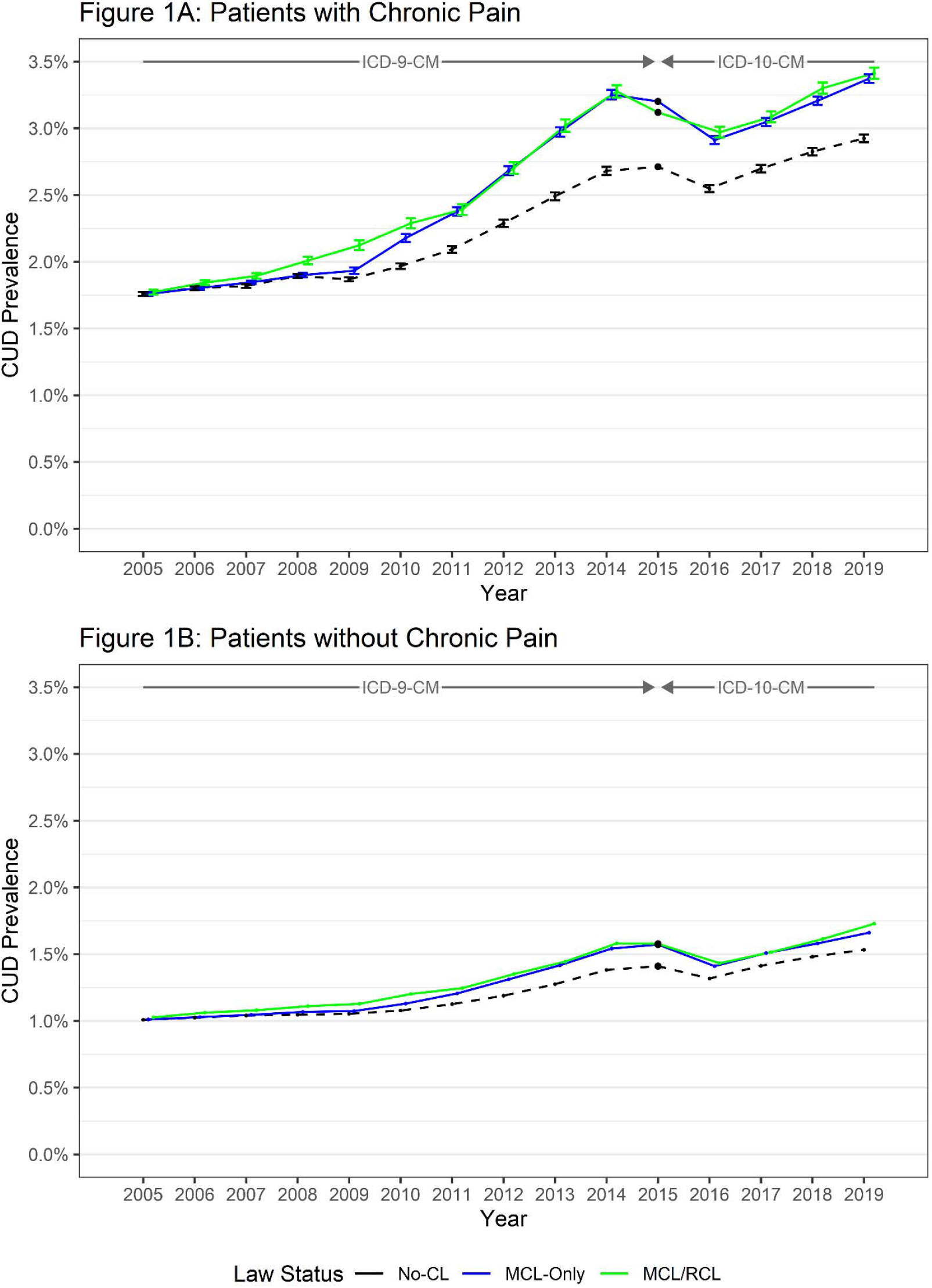
Cannabis Use Disorder prevalence by state law status as of 2019, among patients with and without chronic pain.

In patients with chronic pain by age group, in the no-CL, MCL-only and RCL states, CUD prevalence increased from 2005 to 2019 by 3·64%, 3·89% and 4·12% in patients age 18-34, by 1·92%, 2·57% and 2·45% in patients age 35-64, and by 0·86%, 1·10% and 1·16% in patients age 65-75. In patients without chronic pain, the prevalence increases by state CL status were smaller, ranging from approximately one-third to two-thirds the size of the corresponding increases in patients with chronic pain (Table 1).

### Patients with chronic pain: MCL-and RCL-enactment effects

The DiD estimate of the CUD prevalence increase due to MCL enactment was 0·135% (95%CI 0·118%, 0·153%; Table 2). Relative to the absolute change in states with MCL-only by 2019, 8·4% of this increase was attributable to MCL. The DiD estimate of the CUD prevalence increase due to RCL enactment was 0·188% (95%CI: 0·160%, 0·217%); relative to the absolute change in states with MCL/RCL by 2019, 11·5% of this increase was attributable to RCL.

By age group (Table 2), among those 18-34, neither MCL or RCL played a meaningful or significant role in the increases in CUD prevalence. In patients age 35-64, 7·9% and 5·8% of the CUD prevalence increase was attributable to MCL and RCL; and in patients age 65-75, 9·3% and 19·4% was attributable to MCL and RCL. Thus, in terms of the change attributable to CL, the laws (especially RCL) had greatest impact in the older patients, whose CUD rates were lowest at the beginning of the study period, but increased over 3-fold in MCL or RCL states.

### Patients without chronic pain: MCL- and RCL-enactment effects

The DiD estimate of the CUD prevalence increase due to MCL enactment was 0·037% (95%CI 0·027, 0·048; Table 2). Relative to the absolute change in states with MCL-only by 2019, 5·7% of this increase was attributable to MCL. The DiD estimate of the CUD prevalence increase due to RCL enactment was 0·042%; relative to the absolute change in states with RCL by 2019, 6·0% of this increase was attributable to RCL. By age, MCL and RCL effects were non-significant among patients age 18-34. In patients 35-64, 8·1% of the increase in CUD prevalence was attributable to MCL enactment, while for RCL, the result, -8·2%, suggested that post-RCL enactment, patients age 35-64 without chronic pain had smaller increases in CUD prevalence than others. Among patients age 65-75, 6·9% of the increase in CUD prevalence was attributable to MCL enactment, while 16·1% of the increase was attributable to RCL.

### Patients with and without chronic pain: Comparison of MCL and RCL effects

In the overall sample, a greater increase in CUD prevalence attributable to MCL and RCL occurred among patients with chronic pain, as indicated by the non-overlapping CIs of the DiD estimates for those with and without pain (Table 2). By age, MCL and RCL effects did not differ between those with and without chronic pain in patients age 18-34. However, in patients age 35-64 and 65-75, the non-overlapping CIs for the DiD estimates of MCL and RCL effects indicated that these effects differed significantly between patients with and without pain; MCL and RCL effects were greater in the patients with chronic pain (Table 2).

### State-specific MCL and RCL effects

Figures 2A and 2B show DiD estimates and 95% CIs for patients with and without pain in the states that enacted MCL and/or RCL between 2005-2019, rank-ordered by month/year of enactment. Among patients with pain, 15 states that enacted MCL-only between 2005-2019 showed an increase in CUD prevalence attributable to MCL, 2 a decrease, and 5 no change attributable to MCL; 8 states that enacted RCL by 2019 showed an increase, 1 a decrease and 3 no change attributable to RCL. Among patients without pain, 11 states that enacted MCL-only showed an increase in CUD prevalence attributable to MCL enactment, 8 a decrease, and 3 no change attributable to MCL; 5 states that enacted RCL showed an increase, 2 a decrease and 4 had no change attributable to RCL. Thus, patients with pain had a greater number of state-specific increases attributable to MCL or RCL enactment than other patients. However, no state effect reached a 1% absolute increase in CUD prevalence. We also saw no patterning of larger or smaller effects in states enacting laws in earlier or later years.

**Figure 2.**
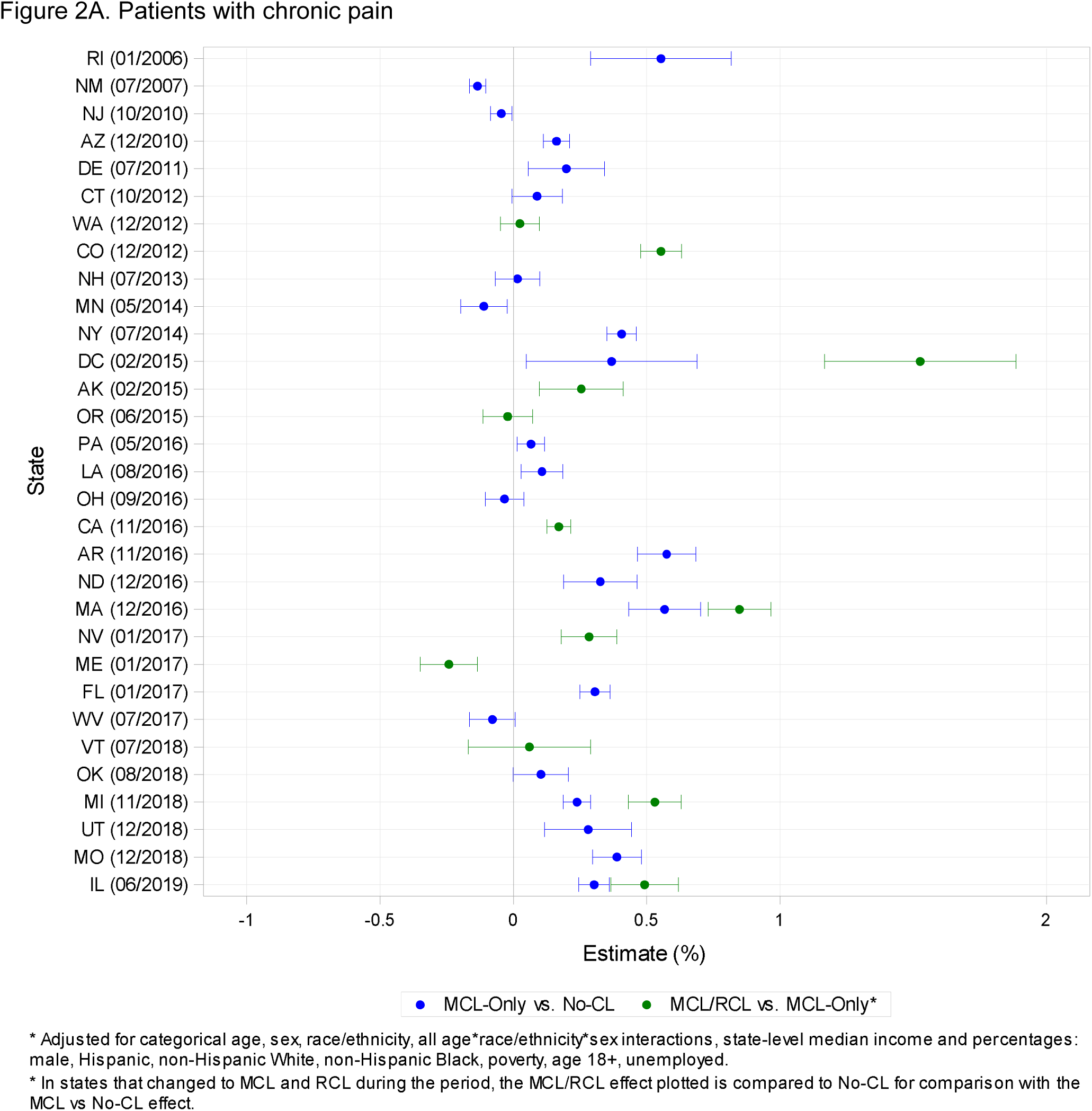
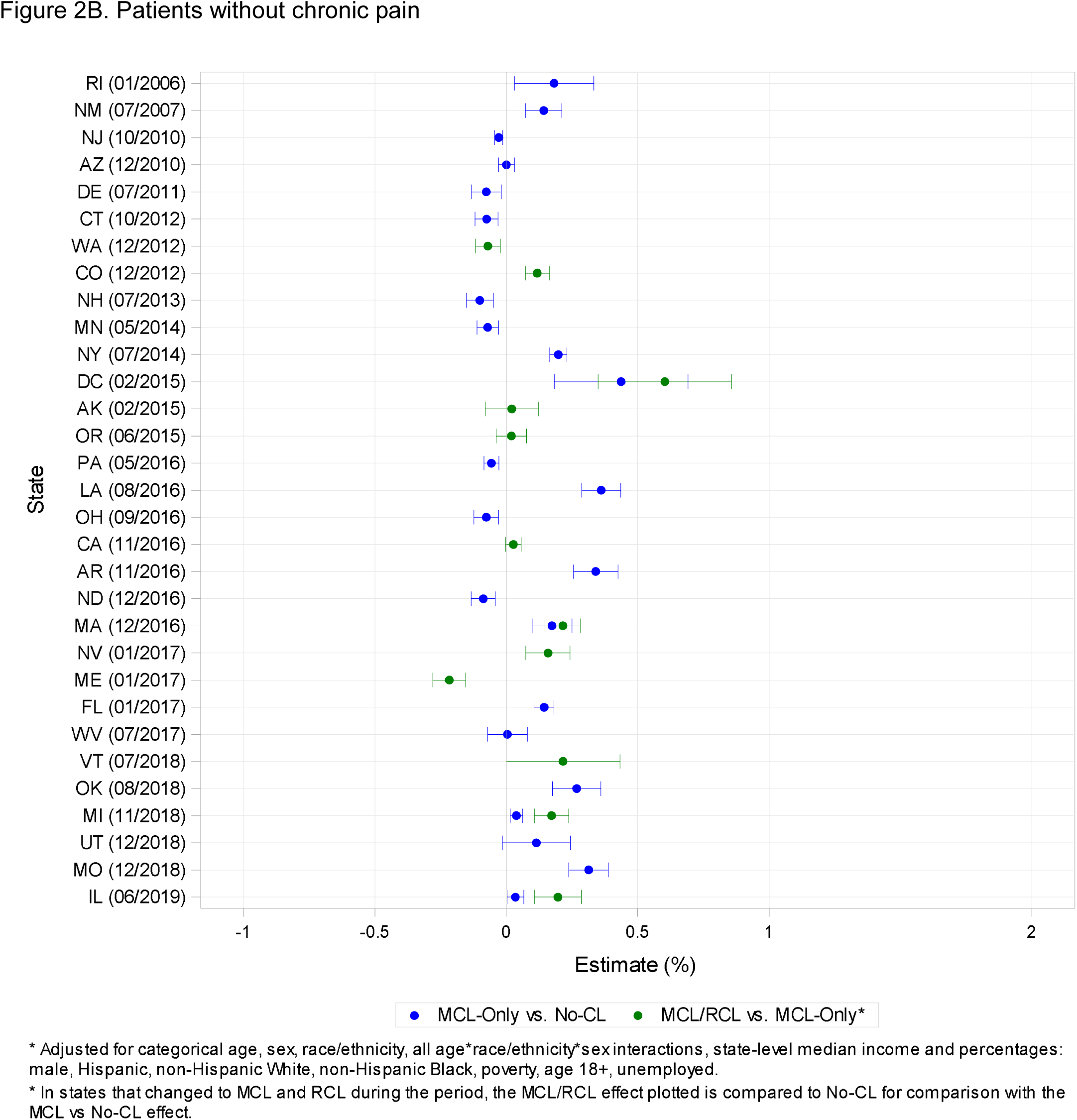
Cannabis Use Disorder Difference-in-Difference estimates, by state: patients with and without chronic pain

### Sensitivity analyses

Substituting legally-permitted operational dispensary dates for law enactment dates (e-Tables 3, 4), fewer states were analyzed because 4 MCL-only and 4 MCL/RCL states had not authorized operational dispensaries by 2019. Using the dates dispensaries were legally permitted among patients with chronic pain, a significant DiD effect was found for MCL (0·040% [95%CI: 0·021, 0·036]) although this effect was smaller than the corresponding result for MCL enactment, and an effect for RCL (0·149% [95%CI: 0·212, 0·182]) that was similar to the effect for the RCL enactment date. Among patients without chronic pain, a small, negative DiD effect was found in MCL-only states (−0·013, 95%CI: -0·023-0·002). Using 1-year lags in place of the dates for MCL and RCL enactment (e-Tables 5, 6), DiD estimates in patients with and without chronic pain did not change substantially.

## DISCUSSION

Chronic pain is a growing problem in the U.S., including among VHA patients, and is associated with CUD. We investigated whether the role of MCL and/or RCL in the increasing prevalence of CUD differed between VHA patients with and without chronic pain. We found that in patients age 35-64 and 65-75, MCL and RCL effects were significantly stronger among patients with chronic pain. In a previous study that did not differentiate VHA patients by pain, MCL and RCL accounted for 4·7% and 9·8% of the 2005-2019 increase in CUD prevalence.^25^ The present study found that among patients with chronic pain, MCL and RCL enactment accounted for 8·4% and 11·5% of the increase in CUD prevalence. RCL effects were particularly pronounced among patients age 65-75 with chronic pain, in whom 19·4% of the increase in CUD prevalence was attributable to RCL enactment.

MCL and RCL are likely to influence the prevalence of CUD through decreased perception of cannabis risk^8, 9^ and commercialization that increases availability and portrays cannabis use as ‘normal’ and safe.^51-54^ In patients with pain, the decline in prescribed opioids^55, 56^ may also have contributed to MCL and RCL effects, leading to substitution of cannabis use that expanded the pool of those vulnerable to CUD. The VHA offers evidence-based non-opioid pain programs.^56^ However, positive media accounts of cannabis trials (regardless of their results^37^), positive online promotional efforts^57^ (e.g., “The Empowered Pain Patient”^58^), and increasing beliefs that cannabis is a useful prevention and treatment agent^36^ may make cannabis more appealing than the evidence-based treatments the VHA offers. That this could increase the risk for CUD is supported by a trial showing that obtaining a medical cannabis card was associated with a rapid increase in risk for CUD,^59^ and also by a meta-analysis of randomized trials of cannabinoids for pain showing substantial placebo response.^37^

Our finding of the largest RCL effects among patients with pain age 65-75 adds to evidence about important changes among older adults, among whom the prevalence of cannabis use is growing.^60, 61^ Many non-opioid pain medications are contraindicated among older individuals,^62, 63^ limiting pain treatment options and potentially leading to cannabis use.^34^ Pharmacokinetic parameters influenced by age may increase tetrahydrocannabinol (THC) bioavailability in older individuals,^64^ possibly increasing their risk for CUD. Taken together, these findings suggest that clinicians should monitor potential cannabis use among older patients with chronic pain, discussing the risks of CUD and potential treatment alternatives.

Although we showed that MCL and RCL played a significant role in the increasing trends in CUD prevalence in patients with chronic pain, a substantial proportion of the increases remains unexplained, leaving important questions unanswered. For example, among patients with chronic pain, are increases in CUD prevalence also influenced by state-level opioid policies or other changing state characteristics? At an individual level, is the prevalence of CUD growing faster among patients with chronic pain who also have specific psychiatric or other substance use disorders, or who were previously prescribed opioids but no longer have a prescription, or who now have a lower dose? Also, in 2019, the two most common chronic pain clusters were back pain (22%) and a category for noninflammatory arthritic pain (32%). Their rates are increasing, as is the use of cannabis in adults with these conditions.^34, 61^ Do one or both of these pain clusters account for most of the increases in CUD prevalence, or for differences in MCL and RCL effects between those with and without chronic pain? Would the total number of painful conditions (a proxy for severity^34, 65, 66^) better account for the increases? While these questions are beyond the scope of this report, they should be addressed in future research.

Study limitations are noted. (1) VHA patients are not representative of all veterans^67-69^ or U.S. adults. (2) CUD diagnoses were made by clinicians, who are most likely to diagnose severe disorders^70, 71^ and miss mild cases found using structured assessments.^7, 10^ While the number of missed cases is unknown and may have varied over time, the overall 2019 VHA CUD prevalence (1·9%)^15^ was similar to the prevalence in a 2019 nationally representative survey (1·7%).^72^ Thus, VHA findings provide a useful counterpart to general population findings with information on what are likely to be severe cases in a large healthcare system where patients cannot all be assessed with research interviews. (3) Cannabis laws are heterogeneous.^73-75^ We examined whether states permitted dispensaries, but other differences (price, taxation) should be addressed in future studies. (4) State law effects may be delayed. We analyzed 1-year lags to include as many RCL states as possible; longer lags should be analyzed as more recent data become available. (5) A cannabis use measure was not available, so CUD within those endorsing use and specific use patterns (e.g., frequency) was not examined. (6) Many time-varying state-level confounders were controlled, but others (e.g., state opioid policies) should be addressed in future studies. (7) The difference-in-differences method estimates law effects in states that enact them, but not spillover effects in adjacent states. If patients in no-CL states are influenced by MCL or RCL laws in nearby states, estimated CL effects will be biased toward the null. For example, patients in no-CL states living near a MCL-only/RCL state^76^ may border-cross to buy cannabis, elevating CUD rates in no-CL states and mitigating MCL or RCL effects. (8) Our chronic pain measure, created from a widely-recognized taxonomy,^77^ was an objective measure. However, no subjective measure (i.e., perceived pain intensity) was available, which future studies should employ. (9) No observational study is unequivocal about causality. However, our pre-post difference-in-difference design controlling for contemporaneous trends and other changing state-level factors provides stronger support for estimated cannabis law effects than studies without such rigor.^78-81^

These limitations are offset by important strengths, including large Ns and rich information from the VHA EHR, enabling us to classify pain according to medically diagnosed conditions. To our knowledge, this is the first study to examine whether MCL and RCL effects on CUD are modified by chronic pain. The study also added to sparse information on an important group, older patients.

Cannabis does not have the same serious overdose/mortality risk as opioids. However, CUD, a disorder with many associated problems,^6, 82-85^ occurs more often among cannabis users than commonly assumed.^4^ The national increases in CUD prevalence, including the disproportionate increase among those with chronic pain, underscores a growing need in the VHA and elsewhere to screen for cannabis use and offer evidence-based treatments for CUD.^86^

The multi-billion-dollar U.S. cannabis industry^87^ requires new customers or greater use among existing customers to increase demand.^51^ Websites of medical cannabis companies often make unconfirmed claims about product safety and efficacy, potentially leading to increasingly positive public beliefs about cannabis use.^52, 54^ Cannabis industry social responsibility programs, ostensibly designed to mitigate harms, mirror tobacco industry programs that actually serve to recruit customers, encourage consumption, expand markets, and legitimize products.^43^ Such activities could change attitudes within and across U.S. state boundaries, increasing cannabis use and CUD, including among those with pain.^37^

In the United States, public health concerns regarding alcohol, tobacco, and prescription opioids have long competed with commercial interests. With cannabis increasingly legalized across U.S. states, similar competing public health and commercial interests are emerging^3^, with state cannabis laws largely industry-friendly rather than protective of public health^3^. In this study, RCL was associated with the greatest increases in CUD prevalence in the oldest patients with chronic pain, who had lower prevalence at the beginning of the study period but greater relative increases by the end. To inform health and policy efforts, researchers should monitor harms related to increasing CUD, particularly in those with chronic pain, and clearly communicate this knowledge to policymakers, clinicians, patients and the public in the U.S. and in other countries.

## Data Availability

No data were collected for this study, which consisted of analyses of Veterans Health Administration (VHA) electronic medical records. These data cannot be made available as they must be kept behind the VHA firewall at all times, and only VHA-authorized users are permitted to access them.

## Author Contributions

Deborah S. Hasin: Conceptualization, Methodology, Software, Validation, Formal analysis, Investigation, Resources, Data curation, Writing – original draft, Writing – review & editing, Visualization, Supervision, Project administration, Funding acquisition; Melanie M. Wall**: Conceptualization, Methodology, Validation, Formal analysis, Writing – original draft, Writing – review & editing, Visualization, Supervision; Dan Alschuler**: Formal analysis, Software, Visualization; Zachary L. Mannes: Methodology, Writing – review & editing; Carol Malte**: Formal analysis, Software, Visualization, Data curation; Mark Olfson: Conceptualization, Methodology, Writing review & editing; Katherine M. Keyes: Katherine M. Keyes: Conceptualization, Methodology, Writing – –review & editing; Jaimie L. Gradus: Conceptualization, Methodology, Writing – review & editing; Magdalena Cerdá: Conceptualization, Methodology, Writing – review & editing; Charles C. Maynard: Methodology, Writing – review & editing; Salomeh Keyhani: Methodology, Writing – review & editing; Silvia S. Martins: Methodology, Writing – review & editing; David S. Fink: Methodology, Writing – review & editing; Ofir Livne: Methodology, Writing – review & editing; Yoanna McDowell: Methodology, Writing – review & editing; Scott Sherman: Writing – review & editing, Project administration; Andrew J. Saxon: Conceptualization, Resources, Writing – review & editing, Supervision, Project administration **Indicates that the author had access to the data.

