## Supplemental Material for "Chronic Pain, Cannabis Legalization and Cannabis Use Disorder in Veterans Health Administration Patients, 2005 to 2019"

**Supplementary Material**

E-Table 1: Patient Ns by year, pain status, and 2019 state cannabis law status for patients included in the prevalence model (page 2)

E-Table 2. Patient Demographics by Any Chronic Pain, 2005 and 2019 (page 3)

E-Table 3. Adjusted CUD prevalence^a^ in VHA patients in 2005 and 2019 by whether states permitted legal operation of dispensaries, and absolute change over time (page 4)

E-Table 4. Effects on CUD prevalence of changes due to whether states legally permitted dispensaries

Difference-in-difference (DiD)^a^ estimates incorporating data on VHA patients across all years 2005 – 2019 (page 5)

E-Table 5. Adjusted CUD prevalence^a^ in VHA patients in 2005 and 2019 by enacted state law status with 1-year lag, and absolute change over time (page 6)

E-Table 6. Effects of changes due to state MCL and RCL enactment plus a 1-year lag on CUD prevalence in VHA patients incorporating data across all years 2005 – 2019: Difference-in-difference (DiD) estimates^a^ (page 7)

| **E-Table 1: Patient Ns by year, pain status, and 2019 state cannabis law status for patients included in the prevalence model.** | | | | | | | | | | | | |
| --- | --- | --- | --- | --- | --- | --- | --- | --- | --- | --- | --- | --- |
|  | **Overall** | | | | **Patients without chronic pain** | | | | **Patients with chronic Pain** | | | |
| **Year** | **Overall** | **None** | **MCL only** | **RCL** | **Overall** | **None** | **MCL only** | **RCL** | **Overall** | **None** | **MCL only** | **RCL** |
| 2005 | 3,234,382 | 1,159,878 | 1,353,409 | 721,095 | 1,952,361 | 683,578 | 818,892 | 449,891 | 1,282,021 | 476,300 | 534,517 | 271,204 |
| 2006 | 3,289,537 | 1,194,583 | 1,361,484 | 733,470 | 1,964,776 | 698,737 | 812,725 | 453,314 | 1,324,761 | 495,846 | 548,759 | 280,156 |
| 2007 | 3,337,837 | 1,220,475 | 1,367,857 | 749,505 | 1,951,815 | 696,731 | 800,081 | 455,003 | 1,386,022 | 523,744 | 567,776 | 294,502 |
| 2008 | 3,404,980 | 1,250,746 | 1,379,758 | 774,476 | 1,943,038 | 698,875 | 789,319 | 454,844 | 1,461,942 | 551,871 | 590,439 | 319,632 |
| 2009 | 3,618,644 | 1,324,601 | 1,460,108 | 833,935 | 2,041,051 | 726,682 | 829,086 | 485,283 | 1,577,593 | 597,919 | 631,022 | 348,652 |
| 2010 | 3,786,314 | 1,388,359 | 1,521,952 | 876,003 | 2,107,740 | 752,466 | 854,396 | 500,878 | 1,678,574 | 635,893 | 667,556 | 375,125 |
| 2011 | 3,942,527 | 1,455,077 | 1,571,263 | 916,187 | 2,194,552 | 793,239 | 876,177 | 525,136 | 1,747,975 | 661,838 | 695,086 | 391,051 |
| 2012 | 4,045,945 | 1,502,718 | 1,603,070 | 940,157 | 2,180,818 | 794,674 | 863,376 | 522,768 | 1,865,127 | 708,044 | 739,694 | 417,389 |
| 2013 | 4,156,005 | 1,545,017 | 1,639,068 | 971,920 | 2,184,067 | 797,652 | 860,689 | 525,726 | 1,971,938 | 747,365 | 778,379 | 446,194 |
| 2014 | 4,303,997 | 1,605,665 | 1,688,419 | 1,009,913 | 2,234,878 | 814,693 | 877,794 | 542,391 | 2,069,119 | 790,972 | 810,625 | 467,522 |
| 2015 | 4,397,541 | 1,650,720 | 1,720,341 | 1,026,480 | 2,230,246 | 815,298 | 873,338 | 541,610 | 2,167,295 | 835,422 | 847,003 | 484,870 |
| 2016 | 4,472,206 | 1,691,419 | 1,741,974 | 1,038,813 | 2,320,109 | 853,242 | 909,172 | 557,695 | 2,152,097 | 838,177 | 832,802 | 481,118 |
| 2017 | 4,519,353 | 1,722,918 | 1,757,122 | 1,039,313 | 2,240,038 | 828,796 | 879,467 | 531,775 | 2,279,315 | 894,122 | 877,655 | 507,538 |
| 2018 | 4,556,541 | 1,745,199 | 1,768,309 | 1,043,033 | 2,158,816 | 800,187 | 845,469 | 513,160 | 2,397,725 | 945,012 | 922,840 | 529,873 |
| 2019 | 4,579,994 | 1,771,798 | 1,775,376 | 1,032,820 | 2,086,807 | 780,943 | 812,351 | 493,513 | 2,493,187 | 990,855 | 963,025 | 539,307 |
| N minima and maxima across 15 years: Overall: min = 3,234,382 (2005), max = 4,579,994 (2019). No pain: min = 1,943,038 (2008), max = 2,320,109 (2016). Pain: min = 1,282,021 (2005), max = 2,493,187 (2019). | | | | | | | | | | | | |

| **E-Table 2. Patient Demographics by Any Chronic Pain, 2005 and 2019** | | | | | | | | | | | | | | |
| --- | --- | --- | --- | --- | --- | --- | --- | --- | --- | --- | --- | --- | --- | --- |
|  | **2005 (N=3,234,382)** | | | | |  | **2019 (N=4,579,994)** | | | | |  | **SMD^a^** | **p^b^** |
|  | **Patients without pain (N=1,952,361)** | |  | **Patients with Pain (N=1,282,021)** | |  | **Patients without pain (N=2,086,807)** | |  | **Patients with Pain (N=2,493,187)** | |  |  |  |
|  | **n** | **% or Mean (SD)** |  | **n** | **% or Mean (SD)** |  | **n** | **% or Mean (SD)** |  | **n** | **% or Mean (SD)** |  |  |  |
| **Age (continuous)** | 1,952,361 | 58·33 (12·59) |  | 1,282,021 | 57.23 (11·39) |  | 2,086,807 | 56·67 (15·17) |  | 2,493,187 | 57·19 (13·78) |  | 0·062 | <0·001 |
| **Age (categorical)** |  |  |  |  |  |  |  |  |  |  |  |  |  | <0·001 |
| **<35** | 122,708 | 6·3% |  | 60,101 | 4·7% |  | 261,075 | 12·5% |  | 221,418 | 8·9% |  | 0·558 |  |
| **35-64** | 1,125,137 | 57·6% |  | 868,355 | 67·7% |  | 939,778 | 45·0% |  | 1,295,657 | 52·0% |  | 0·495 |  |
| **65-75** | 704,516 | 36·1% |  | 353,565 | 27·6% |  | 885,954 | 42·5% |  | 976,112 | 39·2% |  | 0·340 |  |
| **Sex** |  |  |  |  |  |  |  |  |  |  |  |  |  | <0·001 |
| **Female** | 100,228 | 5·1% |  | 90,918 | 7·1% |  | 194,585 | 9·3% |  | 309,659 | 12·4% |  | 0·495 |  |
| **Male** | 1,852,133 | 94·9% |  | 1,191,103 | 92·9% |  | 1,892,222 | 90·7% |  | 2,183,528 | 87·6% |  | 0·495 |  |
| **Race/Ethnicity** |  |  |  |  |  |  |  |  |  |  |  |  |  | <0·001 |
| **White** | 1,477,147 | 75·7% |  | 948,825 | 74·0% |  | 1,420,467 | 68·1% |  | 1,629,004 | 65·3% |  | 0·293 |  |
| **Black** | 305,221 | 15·6% |  | 228,588 | 17·8% |  | 379,810 | 18·2% |  | 546,973 | 21·9% |  | 0·212 |  |
| **Hispanic/Latino** | 70,609 | 3·6% |  | 49,960 | 3·9% |  | 136,680 | 6·5% |  | 174,481 | 7·0% |  | 0·409 |  |
| **Asian** | 11,542 | 0·6% |  | 5,682 | 0·4% |  | 26,669 | 1·3% |  | 28,587 | 1·1% |  | 0·555 |  |
| **Amlnd/AlaskNative** | 10,190 | 0·5% |  | 8,062 | 0·6% |  | 14,597 | 0·7% |  | 19,932 | 0·8% |  | 0·224 |  |
| **PacIs/NatHawaiian** | 13,323 | 0·7% |  | 9,550 | 0·7% |  | 14,052 | 0·7% |  | 19,180 | 0·8% |  | 0·082 |  |
| **MultipleRace/Eth** | 12,175 | 0·6% |  | 10,569 | 0·8% |  | 17,595 | 0·8% |  | 23,717 | 1·0% |  | 0·205 |  |
| **Unknown** | 52,154 | 2·7% |  | 20,785 | 1·6% |  | 76,937 | 3·7% |  | 51,313 | 2·1% |  | 0·452 |  |
| ^a^Standardized absolute mean difference is calculated as the average absolute difference between groups divided by the overall standard deviation. | | | | | | | | | | | | | | |
| ^b^p-values for t-test of continuous variables and chi-square tests of categorical variables. | | | | | | | | | | | | | | |

| **E-Table 3. Adjusted CUD prevalence^a^ in VHA patients in 2005 and 2019 by whether states permitted legal operation of dispensaries, and absolute change over time** | | | | | | |
| --- | --- | --- | --- | --- | --- | --- |
| **Type of State** | **CUD prevalence** | | **Absolute Change** | **CUD prevalence** | | **Absolute Change** |
|  | **2005** | **2019** | **%** | **2005** | **2019** | **%** |
|  | **Patients with chronic Pain** | | | **Patients without chronic pain** | | |
| **No dispensaries by 2019 (17 no-CL, 4 MCL-only states)** | 1·70 | 2·92 | 1·22 | 0·99 | 1·54 | 0·56 |
| **Medical but not recreational dispensary by 2019 (18 MCL-only states, 3 MCL/RCL states and DC)** | 1·71 | 3·38 | 1·67 ^b^ | 0·99 | 1·66 | 0·66^b^ |
| **Recreational dispensary by 2019 (8 MCL/RCL states)** | 1·76 | 3·50 | 1·73^c^ | 1·04 | 1·77 | 0·73^c^ |
| **^a^** Adjusted for categorical age, sex, race/ethnicity, all age*race/ethnicity*sex interactions, state-level median income and percentages: male, Hispanic, non-Hispanic White, non-Hispanic Black, poverty, age 18+, unemployed.  **^b^** Value used as denominator to determine % of overall increase attributable to MCL enactment based on DiD estimates of MCL effect (see E-Table 2)  **^c^** Value used as denominator to determine % of overall increase attributable to RCL enactment based on DiD estimates of RCL effect (see E-Table 2) | | | | | | |

| **E-Table 4. Effects on CUD prevalence of changes due to whether states legally permitted dispensaries**  **Difference-in-difference (DiD)^a^ estimates incorporating data on VHA patients across all years 2005 – 2019** | | | | | |
| --- | --- | --- | --- | --- | --- |
|  | **Model-based DiD law effect**  **%**  **p-value**  **95% CI** | **% of absolute change attributable to law^b^** | **Model-based DiD law effect**  **%**  **p-value**  **95% CI** | **% of absolute change attributable to law^b^** | **Difference in effect between Pain and no Pain** |
|  | **Patients with chronic pain** | | **Patients without chronic pain** | |  |
| **Effect of change to medical dispensaries^c^** | 0·040 (<·001) 0·021, 0·059 | 2·4% | -0·013 (0·016) -0·023, -0·002 | -1·9% | <0·01 |
| **Effect of change to recreational dispensaries^c^** | 0·149 (<·001) 0·115, 0·182 | 8·6% | 0·069 (<·001) 0·047, 0·092 | 9·5% | <0·01 |
| ^a^ Staggered-Adoption Difference-in-Difference (DID) regression model(88), adjusted for categorical age, sex, race/ethnicity, all age*race/ethnicity*sex interactions, state-level median income and percentages: male, Hispanic, non-Hispanic White, non-Hispanic Black, poverty, age 18+, unemployed. Model estimated effects represent absolute increase (positive values) or decrease (negative values) in CUD prevalence due to law enactment. Confidence intervals not including 0·0 indicate significant changes. The DiD model compares the years after enactment (up to 2019 or until the next law change) in each state to the years before enactment (since 2005 or the previous law change) in the same state, and controls for contemporaneous trends in other states that have not yet passed the respective law.  ^b^ DiD estimate divided by absolute change across period as shown in E-Table 1.  ^c^ 21 states and DC made a change to having medical dispensaries during the period; 2 states made a change to having recreational dispensaries and 6 states made a change to medical and to recreational dispensaries | | | | | |

| **E-Table 5. Adjusted CUD prevalence^a^** **in VHA patients in 2005 and 2019 by enacted state law status with 1-year lag, and absolute change over time** | | | | | | |
| --- | --- | --- | --- | --- | --- | --- |
| **Type of State** | **CUD prevalence**  **%** | | **Absolute Change** | **CUD prevalence**  **%** | | **Absolute Change** |
|  | **2005** | **2019** | **%** | **2005** | **2019** | **%** |
|  | **Patients with chronic pain** | | | **Patients without chronic pain** | | |
| **No-CL (17 states by 2019)** | 1·73 | 2·92 | 1·19 | 1·00 | 1·53 | 0·54 |
| **MCL-Only (23 states by 2019)^d^** | 1·74 | 3·38 | 1·64^b^ | 1·00 | 1·67 | 0·67^b^ |
| **MCL/RCL (10 states and DC by 2019)^d^** | 1·77 | 3·44 | 1·67^c^ | 1·03 | 1·75 | 0·71^c^ |
| ^a^ Adjusted for categorical age, sex, race/ethnicity, all age*race/ethnicity*sex interactions, state-level median income and percentages: male, Hispanic, non-Hispanic White, non-Hispanic Black, poverty, age 18+, unemployed.  **^b^** Value used as denominator to determine % of overall increase attributable to MCL enactment based on DiD estimates of MCL effect (see E-Table 4)  **^c^** Value used as denominator to determine % of overall increase attributable to RCL enactment based on DiD estimates of RCL effect (see E-Table 4)  ^d^ MCL-only and MCL/RCL states differ from Table 2 by 1 because Illinois (IL) which enacted RCL in 2019 is considered a MCL-only state here to account for the 1-year lag | | | | | | |

| **E-Table 6. Effects of changes due to state MCL and RCL enactment plus a 1-year lag on CUD prevalence in VHA patients incorporating data across all years 2005 – 2019: Difference-in-difference (DiD) estimates^a^** | | | | | |
| --- | --- | --- | --- | --- | --- |
|  | **Model-based DiD law effect**  **%**  **p-value**  **95% CI** | **% of absolute change attributable to law^b^** | **Model-based DiD law effect**  **%**  **p-value**  **95% CI** | **% of absolute change attributable to law^b^** | **Difference in effect between Pain and no Pain** |
|  | **Patients with chronic Pain** | | **Patients without chronic pain** | |  |
| **Effect of change from no-CL to MCL-only^c^** | 0·124 (<·001) 0·105, 0·143 | 7·6% | -0·012 (0·045) -0·023, -0·000 | -1·8% | <0·01 |
| **Effect of change from MCL-only to RCL^c^** | 0·240 (<·001) 0·207, 0·273 | 14·4% | 0·077 (<·001) 0·056, 0·098 | 10·8% | <0·01 |
| ^a^ Staggered-Adoption Difference-in-Difference (DID) regression model(88), adjusted for categorical age, sex, race/ethnicity, all age*race/ethnicity*sex interactions, state-level median income and percentages: male, Hispanic, non-Hispanic White, non-Hispanic Black, poverty, age 18+, unemployed.  Model estimated effects represent absolute increase (positive values) or decrease (negative values) in CUD prevalence due to law enactment. Confidence intervals not including 0·0 indicate significant changes. The DiD model compares the years after enactment (up to 2019 or until the next law change) in each state to the years before enactment (since 2005 or the previous law change) in the same state, and controls for contemporaneous trends in other states that have not yet passed the respective law.  ^b^ DiD estimate divided by absolute change across period as shown in E-Table 3.  ^c^ 22 states and DC made a change from no-CL to MCL-only during the period from 2005-2018 (to account for the 1-year lag); 10 states and DC made a change from MCL-only to RCL/MCL during the period. Note, 2 of these states and DC made both changes during the period from no-CL to MCL-only and then to RCL/MCL hence contribute to both effects. With the 1-year lag, 20 states (3 with MCL-only and 17 with no-CL) made no law changes between 2005-2019 and contribute in the DID model to background secular trends. | | | | | |
